# Smart youth: sociodemographic factors, usage patterns, and self-reported vs. actual smartphone addiction among secondary school students

**DOI:** 10.1101/2024.04.17.24305981

**Authors:** Magdalena Rękas, Joanna Burzyńska

## Abstract

**Background:** Smartphone addiction is a growing social problem especially in young mobile users. This study investigated indicators of smartphone use, smartphone addiction, and their associations with demographic and behavior-related variables in young people.

**Methods:** 460 participants were secondary school students (M_age_ = 17,10, SD_age_ = 0.92, 51.1% males, 52.4% high school students), took part in an anonymous questionnaire consisting of the following elements: the Mobile Phone Addiction Assessment Questionnaire (KBUTK), original questions regarding problematic smartphones usage, along with a subjective assessment of the use of such devices. Logistic regression model using forward stepwise method was used to characterize a typical smartphone user. Smartphone addiction was measured using KBUTK. Multiple logistic regression analysis was performed to determine factors associated with smartphone addiction.

**Results:** A total of 460 participants admitted to using a smartphone. Gender, age, type of school, place of living influenced the ways respondents used their smartphones. Being female (OR = 5.80; *p* < 0.0001), sixteen-year-old (OR = 0,41; *p* = 0.0456), and student of technical school (OR = 2.66; *p* = 0.0025) turned out to be the characteristics of a typical smartphone user. 21.7% of adolescents considered themselves addicted to smartphones, 22.2% admitted that they had problems with face-to-face relationships and girls significantly more often than boys (61.8% vs. 51.5%) neglected home or school duties as a result of using a smartphone. The overall rate of smartphone addiction was significantly higher (*p* < 0.0001) among girls (2.31 pts) than boys (2.03 pts), and correlated positively with the perception of being a smartphone addict (*rho* = 0.223; *p* < 0.0001). Addiction to smartphones was also significantly more common among students of technical schools, and respondents living in blocks of flats.

**Conclusions:** The way adolescents used smartphones differed depending on gender, age and type of school. Interventions for reducing the negative effects of smartphone use should take into account these context, as well as education both adolescents and their parents.

## Introduction

In an era where digital devices have become ubiquitous, adolescents are at the forefront of embracing smartphone technology. These pocket-sized gadgets serve as portals to a vast digital universe, offering connectivity, entertainment, and information at the swipe of a finger. While smartphones undoubtedly bring numerous benefits, concerns have arisen regarding their potential impact on adolescent development, particularly concerning excessive use and its consequences.

There is growing evidence that smartphones are being overused in ways that are forcing changes in their users’ daily lives and health [1, 2]. It has been shown that the frequency of smartphone use, especially by adolescents, may be associated with negative effects on mental health, such as depression [2–5], anxiety [2–4], and addiction [6–8]. In literature and colloquial discourse, smartphone addiction is defined by various terms, and might generate completely new dangers for users. According to the Diagnostic and Statistical Manual of Mental Disorders - DSM IV addiction is “a mental or physical compulsion to perform certain activities or take certain substances in anticipation of their effects or to avoid the unpleasant symptoms of their absence” [9]. The phenomenon associated with smartphone addiction is phonoholism, defined as an excessive and uncontrolled use of mobile phone/smartphone functions. Phonoholic has a strong need to keep the device close and has problems with turning it off or not using it in situations where it is not necessary. It might be the reason of various ailments, such as dizziness, shortness of breath, accelerated heartbeat, nausea, abdominal pain or headaches [10].

Before addiction occurs, overuse and problematic smartphone use is common. Problematic usage has been linked to “fear of missing out” syndrome (FoMo), described as a state of mind in which smartphone users experience anxiety associated with separation from their device [11–13]. It has been shown that the reactions of FoMo and anxiety related to the frequency of smartphone use are characteristic of attachment theory, known as: motivation to be with the attachment figure/object and anxiety when it is absent [14]. It has also been shown that smartphone use can affect sleep patterns and reduce sleep quality [10, 15, 16]. In a 14-day, randomized, crossover experimental study under well-controlled conditions, the use of electronic screens before sleep was shown to disrupt sleep in many ways: it increases the time to fall asleep and reduces evening sleepiness, reduces melatonin secretion, delays the circadian clock, and reduces next-day alertness [15]. In turn, a study of American teenagers found that spending more than a few hours a week using electronic media was negatively correlated with feelings of happiness, life satisfaction and self-esteem, while time spent on non-screen activities (personal interactions, sports or exercise, etc.) positively correlated with the mental well-being of the surveyed youth [16]. Apart from communication and entertainment functions, smartphones also serve as identity functions which manifests itself in setting an individual wallpaper or selecting a case or the type of ringtone. This mobile personalization refers to the degree to which users customize the device to express themselves through both the appearance of the device and its settings [17]. Modern mobile devices allow not only to establish or maintain contacts thanks to voice and video calls, but also constitute personal elements, often being an expression of prestige and personality [18]. The above features are particularly important during adolescence.

Based on these considerations, the objectives of the study were to evaluate the sociodemographic factors and usage patterns of smartphones while identifying the differences between self-assessments of the use of this type of devices with objectively measured data among adolescents. By examining the disparities between self-perception and actual usage, we can shed light on the reliability of self-reports in assessing smartphone usage accurately. Additionally, exploring factors influencing these disparities can provide valuable insights into adolescents’ perceptions and behaviors regarding smartphone use.

### Characteristics of the smartphone market

In 2012, 9% of Poles were aware that they used a smartphone, while in fact every fourth person had one [19]. At that time, users did not realize the difference between a regular mobile phone and a smartphone. A smartphone combines the functions of a mobile phone and a portable computer, and the development of new technologies forces consumers to search for better and more functional devices [20]. In Poland, smartphone use is progressing at a dynamic pace. According to “Poland in numbers 2019” report, smartphone is a dominant tool for accessing Internet in each age group [19]. One fourth of respondents declare that they send and receive e-mails using the device and 23% of respondents make mobile shopping, and 13% of respondents make payments by phone [19].

According to the Global Digital 2023 report, over two-thirds (68%) of the world’s population now use a mobile phone, and the number of unique mobile users has increased year-on-year by just over 3 percent, reaching 168 million new users in the last 12 months [21]. There are 851 million smartphone owners in China, which is the largest number of users and translates into 59.9% of the population living in this country, while in Switzerland there are 6.2 million smartphone users, which is 72.9% of the population [21]. The United Kingdom has the highest ratio of smartphone users to population in a given country. As many as 82.9% of English citizens have a smartphone, which is 55.5 million users [21]. There are also clear global disproportions, e.g. the population of smartphone owners in Nigeria is less than 15% [21]. In the United States, the percentage of 13- to 17-year-olds who own a smartphone has reached 89%, more than doubling in 6 years [21]. According to the findings of the Office of Electronic Communications, over 80% of Polish children aged 7 to 15 have a mobile phone, while in the age group 13 to 15, each child has their own phone [22]. More than 90% of it is a smartphone. Over 70% of children declare that they use smartphones to play and listen to music, and in over 60% of cases to browse websites [22]. Unfortunately, only less than half of parents control their child’s use of a smartphone. Reports from research conducted among parents of children aged 6 months to 6.5 years showed that 64% of children use mobile devices [23]. According to parents of children aged between 5 and 6, only 17% did not report such use [23]. Polish nationwide research conducted among high school students confirmed that 18% of respondents believe that they spend too much time using a smartphone, and 17% believe that they do not use a smartphone very often, even though others point this out [24]. 32% of young respondents admitted that they spend 4 to 5 hours a day with their smartphone. Boys use handheld devices longer than girls by about 15 minutes [24].

## Materials and methods

### Ethic statement

The study was approved by the institutional Bioethics Committee of the Rzeszow University – Resolution No. 28/02/2019. Moreover, consent from the management to conduct the research was obtained from the secondary schools participating in the project. During meetings with parents/legal guardians, written consent was obtained. The respondents themselves gave verbal consent to participate in the study.

### Participants

The sample consisted of secondary school students (N = 460) from the south-eastern region of Poland (Podkarpackie Voivodeship) aged 16-19 years old, recruited from September to December 2019. There were: 235 (51.1%) boys and 225 (48.9%) girls. The average age of the respondents was 17.10±0.92 years. More than half of the youth (N = 241, 52.4%) attended high school.

### Smartphone use

Respondents were asked about the following issues related to the use of a smartphone: a) owning a smartphone; b) age of receiving the first smartphone; c) features that determine the purchase of the device; and d) ways of using the smartphone.

### Subjective variables associated with problematic smartphone use

Participants were also asked about subjective variables associated with problematic smartphone such as: a) respondents’ opinion about the possible addiction; b) problems with establishing face-to-face contacts; c) neglecting home/school duties; d) inability to spend time without a smartphone; e) returning home in case forgetting the device; f) situations when the smartphone is not used, and g) respondents’ opinion about smartphone usage and health status.

### The degree of smartphone addiction

The degree of smartphone addiction was assessed using the Mobile Phone Addiction Assessment Questionnaire (KBUTK) by Pawłowska and Potembska [25]. Written consent from the authors was obtained to use the tool. KBUTK consists of 33 items that are rated on a five-point Likert scale, where zero (0) means “never” and four (4) means “always”. The questionnaire is applied to examine addiction to mobile phone in the four dimensions: 1) “Need of acceptance and closeness”, 2) “Addiction to camera function”, 3) “Addiction to phone calls and text messages”, and 4) “Intermediary communication”. The results may range from 0 to 132 points. Respondents who obtained a score from 31 to 69 were considered to be at risk of smartphone addiction, and those who scored 70 or more points were considered addicted [25].

### Statistical analyses

Descriptive statistics (frequencies, percentages, means, and standard deviations) were used to examine the gender and age of each participant, place of residence, type of school, living conditions, and family economic condition. When assessing the differences between two nominal variables the *chi^2^* test of independence was used, taking into account the Yates correction. The selection of tests to assess differences between quantitative and nominal variables was made after previously assessing the normality of variable distributions using the Kolmogorov-Smirnov test. The lack of normality of variable distributions suggested the use of Spearman’s *rho* correlation coefficient. Cronbach’s α coefficient was used to assess the reliability of KBUTK scale. Logistic regression model using forward stepwise method (likelihood ratio) was used to characterize a typical smartphone user in this study. Logistic regression models using input method or forward selection method were used to assess the significance of selected correlates of the dependent variable “smartphone addiction”. *P*-values lower than 0.05 were considered significant. All the statistical analyses were performed using SPSS Statistics (version 20.0, IBM Corp., Armonk, NY, USA). Adjusted ORs (ORs) and 95% confidence intervals (CIs) were calculated.

## Results

### Smartphone use

#### Owning the device

All respondents admitted that they use a smartphone. Almost all participants (N = 445; 96.7%) have such a device for they own. Table 1 shows that these were more often girls than boys (98.7% vs. 94.9%; *χ^2^* = 4.060; *p* = 0.0439). Age of the respondents did not significantly affect this issue (*χ^2^* = 4.729; *p* = 0.0940).

**Table 1.**
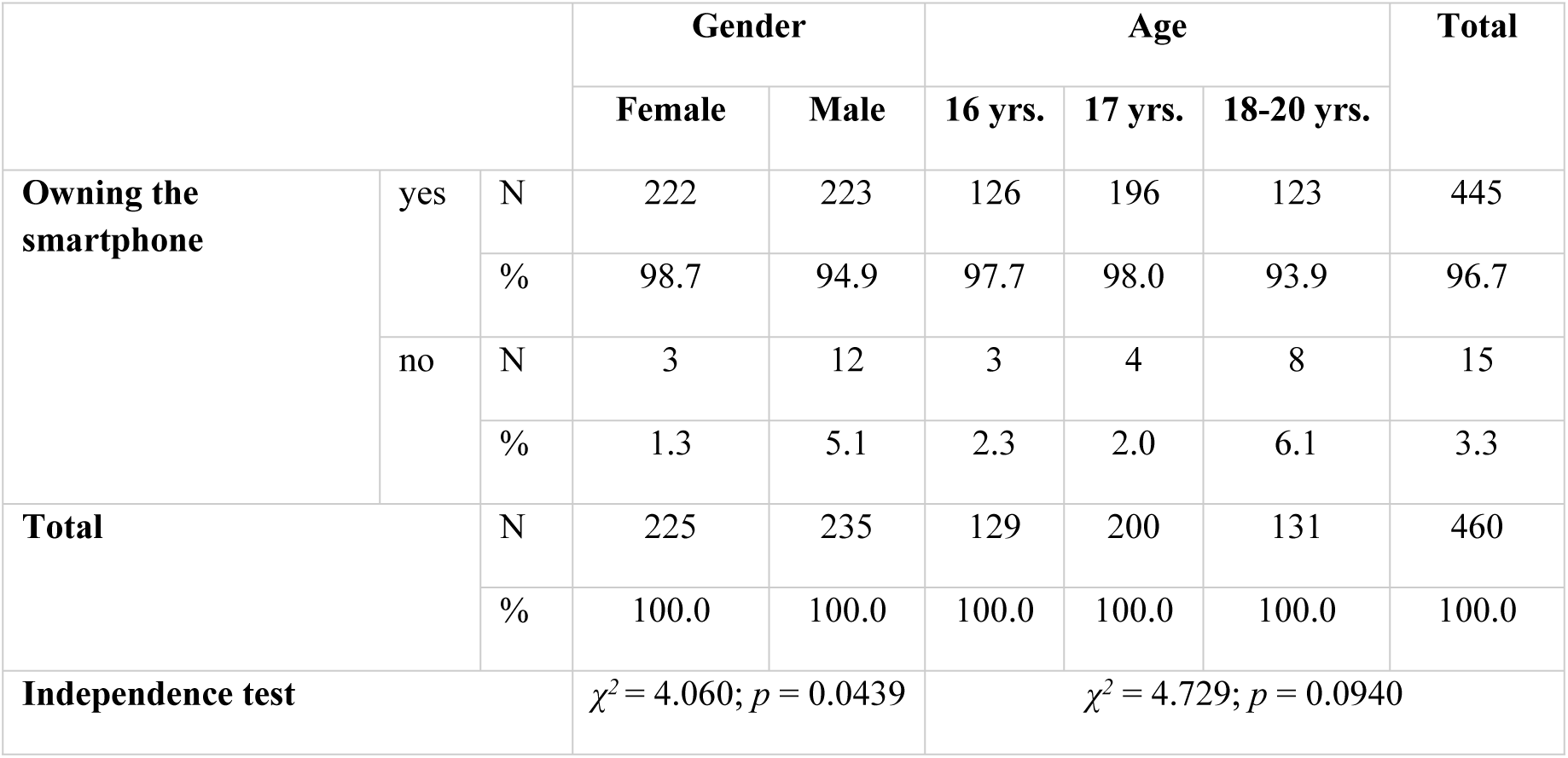
Owning a smartphone and gender and age.

#### Age of receiving the first smartphone

We have asked participants when they received their first device. 18.3% (N = 84) of students admitted they were under 10 years old when they received their first smartphone. Most often, respondents received their first mobile device at the age of 10-13 (N = 283; 61.5%). Every fifth person (N = 93; 20.2%) received their first smartphone when they were between 14 and 16 years old. Girls were more likely than boys (24.0% vs. 12.8%) to receive their first smartphone under the age of 10; boys, however, were more likely to receive their first device at the age of 14-16 (24.7% vs. 15.6%). Gender influence significantly the age of receiving the first smartphone (*χ^2^* = 12.762; *p* = 0.0017). It was noticed that the age of the respondents also significantly influenced the stage of life in which they received first mobile device (*χ^2^* = 19.029; *p* = 0.0008). At the age of 10-13, current 16-year-olds (N = 147; 69.8%) or 17-year-olds (N = 90; 62.0%) received their first smartphone more often. Less often respondents who were or had reached the age of majority (N = 69; 52.7%). The analysis also showed that high school students received their first smartphone more often under the age of 10 (N = 60; 24.9%) or at the age of 10-13 (N = 157; 65.1%), and this differences were statistically significant (*χ^2^* = 39.637; *p* < 0.0001). Family financial situation of the respondents had a statistically significant relationship (*χ^2^* = 19.998; *p* < 0.0001) with the age of receiving first smartphone. According to the analyses, under the age of 10, such a device was more often received by respondents whose family financial situation was good (N = 65; 25.1%) compared to those with an average financial situation (25.1% vs. 9.5%). Later in life, the first smartphone was received by adolescents with an average financial situation in their family (24.9% vs. 16.6%). Living conditions and place of residence did not significantly affect the age at which young people received their first smart phone – Table 2.

**Table 2.** Respondents’ age of receiving the first smartphone and sociodemographic variables.

|  |  | Age of receiving the first smartphone |  |  |  |  |  | Independence test |
| --- | --- | --- | --- | --- | --- | --- | --- | --- |
|  |  | under 10 yrs. |  | 10-13 yrs. |  | 14-16 yrs. |  |  |
|  |  | N | % | N | % | N | % |  |
| Gender | Female | 54 | 24.0 | 136 | 60.4 | 35 | 15.6 | $\chi^2 = 12.762$; |
| | Male | 30 | 12.8 | 147 | 62.6 | 58 | 24.7 | $p = 0.0017$ |
| Age | 16 yrs | 17 | 13.2 | 90 | 69.8 | 22 | 17.1 | $\chi^2 = 19.029$; |
| | 17 yrs | 46 | 23.0 | 124 | 62.0 | 30 | 15.0 | $p = 0.0008$ |
|  | 18-20 yrs | 21 | 16.0 | 69 | 52.7 | 41 | 31.3 |  |
| Type of school | High school | 60 | 24.9 | 157 | 65.1 | 24 | 10.0 | $\chi^2 = 39.637$; |
| | Technical school | 24 | 11.0 | 126 | 57.5 | 69 | 31.5 | $p < 0.0001$ |
| Place of residence | City | 30 | 24.6 | 70 | 57.4 | 22 | 18.0 | $\chi^2 = 4.498$; |
| | Village | 54 | 16.0 | 213 | 63.0 | 71 | 21.0 | $p = 0.1055$ |
| Living conditions | House | 74 | 18.4 | 250 | 62.2 | 78 | 19.4 | $\chi^2 = 1.315$; |
| | Flat | 10 | 17.2 | 33 | 56.9 | 15 | 25.9 | $p = 0.5183$ |
| Family economic | Average | 19 | 9.5 | 132 | 65.7 | 50 | 24.9 | $\chi^2 = 19.998$; |
| condition | Good | 65 | 25.1 | 151 | 58.3 | 43 | 16.6 | $p < 0.0001$ |

#### The main features that determine the purchase of a smartphone

When deciding to buy a smartphone, the surveyed youth were most often guided by battery life (N = 280; 60.9%), technical parameters (N = 273; 59.3%), price (N = 272; 59.1%), and having a good camera on the smartphone (N = 269; 58.5%). 44.8% of students (N = 206) paid attention to the brand. More than every third respondent (N = 161; 35.0%) paid attention to the size of the device, and every third (N = 153; 33.3%) was interested in the possibility of installing various applications. Every fourth student was guided by its colour (N = 117; 25.4%). Examined youth rarely took into account that the device was fashionable (N = 53; 11.5%), easy to use (N = 31; 6.7%), and that a friend had such an equipment (N = 20; 4.3%) or other arguments (N = 4; 0.9%) – Fig 1.

**Fig. 1.**
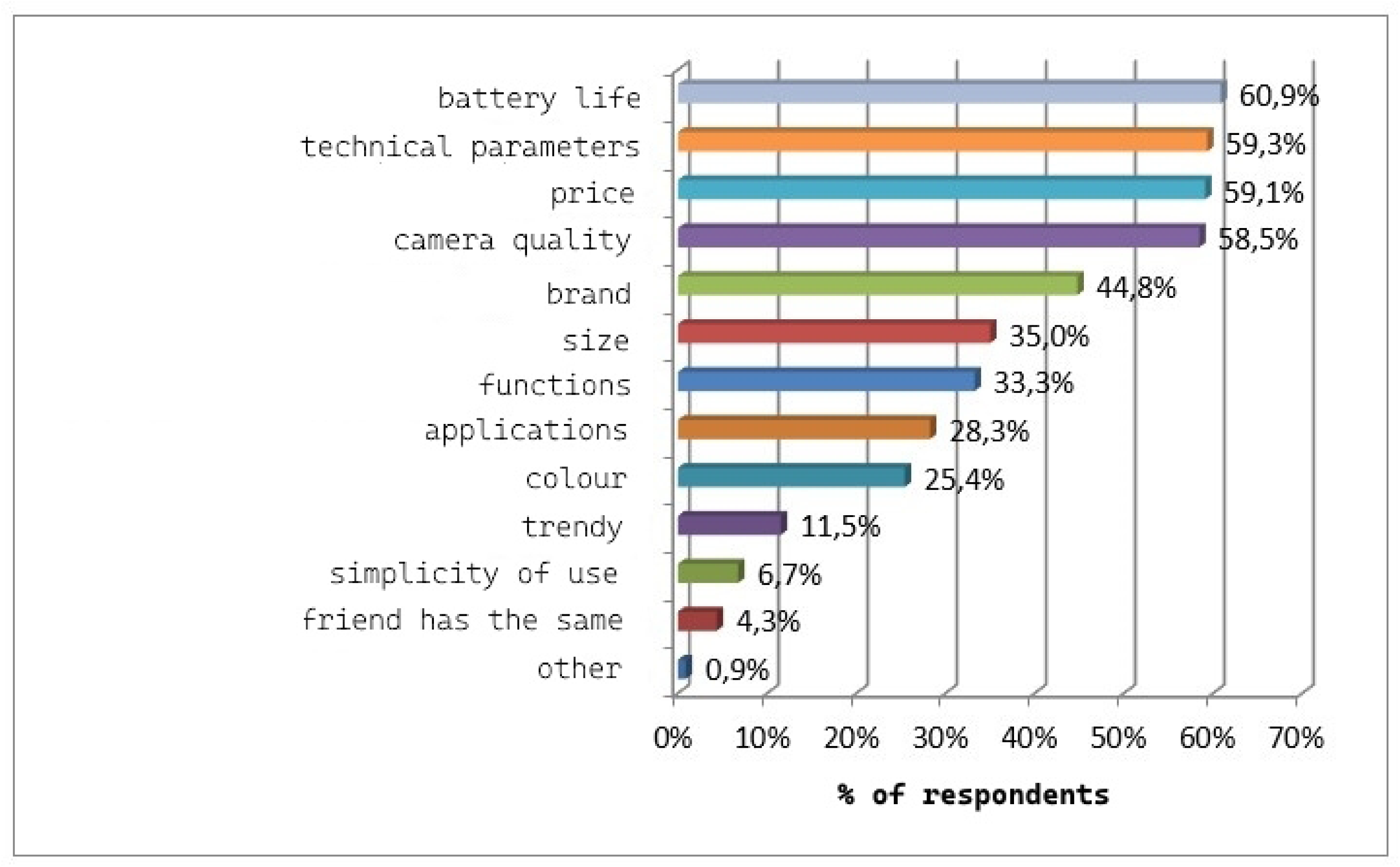
Features that determine the purchase of a new smartphone. The results did not sum up to 100% - multiple choice question.

Table 3 showed that when deciding to buy a new smartphone, girls were significantly more likely than boys to be guided by the price of the product (66.7% vs. 51.9%; *χ^2^* = 10.351; *p* = 0.0013), having a good camera (74.7% vs. 43.0%; *χ^2^* = 47.535; *p* < 0.0001), colour (33.3% vs. 17.9%; *χ^2^* = 14.488; *p* = 0.0001), and the possibility of installing various applications (37.3 % vs. 19.6%; *χ^2^* = 17.881; *p* < 0.0001). Boys in turn were more likely than girls to pay attention to the device’s technical parameters (69.4% vs. 48.9%; *χ^2^* = 19.969; *p* < 0.0001). It was also noticed that 16-year-olds were more likely (10.9%) than older students to be guided by the fact that a friend had an identical device (*χ^2^* = 18.359; *p* = 0.0001). When purchasing a smartphone among high school students price (65.1% vs. 52.5%; *χ^2^* = 7.578; *p* = 0.0059), having a good camera (68.0% vs. 47.9; *χ^2^* = 6.828; *p* < 0.0001), colour (32.0% vs. 18.3%; *χ^2^*=11.331; *p* = 0.0008), and the ability to install various applications (32.8% vs. 23.3%; *χ2* = 5.099; *p* = 0.0239) mattered. Students of technical schools paid more attention to the size (41.1% vs. 29.5%; *χ^2^* = 6.828; *p* = 0.0090), and simplicity of use (11.0% vs. 2.9%; *χ^2^* = 11.843; *p* = 0.0006) compared to high school students. It was also noticed that urban residents paid attention to the brand more often than rural residents when purchasing a smartphone (54.9% vs. 41.1%; *χ^2^* = 6.898; *p* = 0.0086).

**Table 3.**
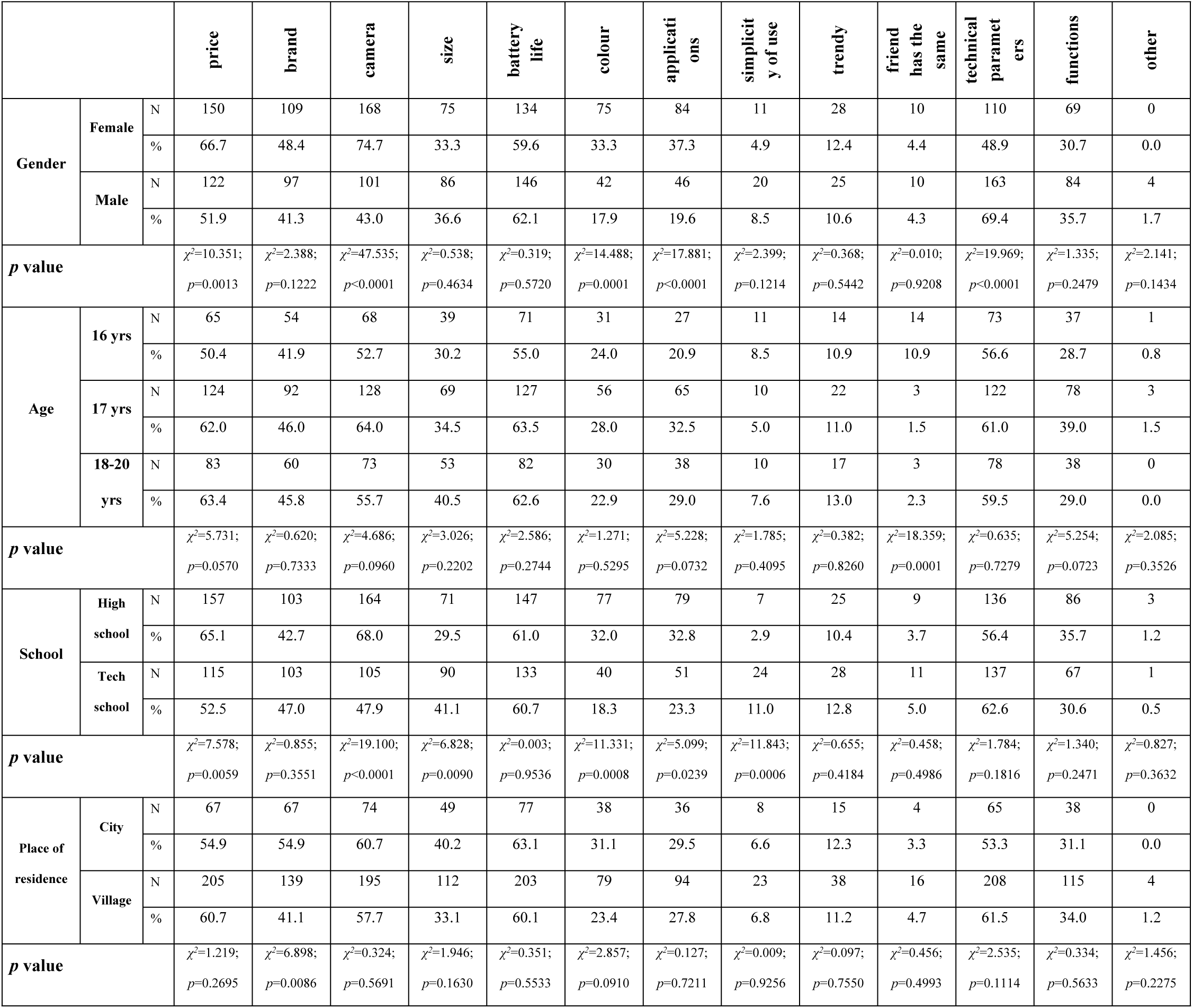
Features that determine the purchase of a new smartphone and sociodemographic variables.

|  |  |  | price | brand | camera | size | battery<br>life | colour | applicati<br>ons | simplicit<br>y of use | trendy | friend<br>has the<br>same | technical<br>paramet<br>ers | functions | other |
| --- | --- | --- | --- | --- | --- | --- | --- | --- | --- | --- | --- | --- | --- | --- | --- |
| Gender | Female | N | 150 | 109 | 168 | 75 | 134 | 75 | 84 | 11 | 28 | 10 | 110 | 69 | 0 |
|  |  | % | 66.7 | 48.4 | 74.7 | 33.3 | 59.6 | 33.3 | 37.3 | 4.9 | 12.4 | 4.4 | 48.9 | 30.7 | 0.0 |
|  | Male | N | 122 | 97 | 101 | 86 | 146 | 42 | 46 | 20 | 25 | 10 | 163 | 84 | 4 |
|  |  | % | 51.9 | 41.3 | 43.0 | 36.6 | 62.1 | 17.9 | 19.6 | 8.5 | 10.6 | 4.3 | 69.4 | 35.7 | 1.7 |
| p value | | | $\chi^2=10.351$;<br>$p=0.0013$ | $\chi^2=2.388$;<br>$p=0.1222$ | $\chi^2=47.535$;<br>$p<0.0001$ | $\chi^2=0.538$;<br>$p=0.4634$ | $\chi^2=0.319$;<br>$p=0.5720$ | $\chi^2=14.488$;<br>$p=0.0001$ | $\chi^2=17.881$;<br>$p<0.0001$ | $\chi^2=2.399$;<br>$p=0.1214$ | $\chi^2=0.368$;<br>$p=0.5442$ | $\chi^2=0.010$;<br>$p=0.9208$ | $\chi^2=19.969$;<br>$p<0.0001$ | $\chi^2=1.335$;<br>$p=0.2479$ | $\chi^2=2.141$;<br>$p=0.1434$ |
| Age | 16 yrs | N | 65 | 54 | 68 | 39 | 71 | 31 | 27 | 11 | 14 | 14 | 73 | 37 | 1 |
|  |  | % | 50.4 | 41.9 | 52.7 | 30.2 | 55.0 | 24.0 | 20.9 | 8.5 | 10.9 | 10.9 | 56.6 | 28.7 | 0.8 |
|  | 17 yrs | N | 124 | 92 | 128 | 69 | 127 | 56 | 65 | 10 | 22 | 3 | 122 | 78 | 3 |
|  |  | % | 62.0 | 46.0 | 64.0 | 34.5 | 63.5 | 28.0 | 32.5 | 5.0 | 11.0 | 1.5 | 61.0 | 39.0 | 1.5 |
|  | 18-20<br>yrs | N | 83 | 60 | 73 | 53 | 82 | 30 | 38 | 10 | 17 | 3 | 78 | 38 | 0 |
|  |  | % | 63.4 | 45.8 | 55.7 | 40.5 | 62.6 | 22.9 | 29.0 | 7.6 | 13.0 | 2.3 | 59.5 | 29.0 | 0.0 |
| p value | | | $\chi^2=5.731$;<br>$p=0.0570$ | $\chi^2=0.620$;<br>$p=0.7333$ | $\chi^2=4.686$;<br>$p=0.0960$ | $\chi^2=3.026$;<br>$p=0.2202$ | $\chi^2=2.586$;<br>$p=0.2744$ | $\chi^2=1.271$;<br>$p=0.5295$ | $\chi^2=5.228$;<br>$p=0.0732$ | $\chi^2=1.785$;<br>$p=0.4095$ | $\chi^2=0.382$;<br>$p=0.8260$ | $\chi^2=18.359$;<br>$p=0.0001$ | $\chi^2=0.635$;<br>$p=0.7279$ | $\chi^2=5.254$;<br>$p=0.0723$ | $\chi^2=2.085$;<br>$p=0.3526$ |
| School | High<br>school | N | 157 | 103 | 164 | 71 | 147 | 77 | 79 | 7 | 25 | 9 | 136 | 86 | 3 |
|  |  | % | 65.1 | 42.7 | 68.0 | 29.5 | 61.0 | 32.0 | 32.8 | 2.9 | 10.4 | 3.7 | 56.4 | 35.7 | 1.2 |
|  | Tech<br>school | N | 115 | 103 | 105 | 90 | 133 | 40 | 51 | 24 | 28 | 11 | 137 | 67 | 1 |
|  |  | % | 52.5 | 47.0 | 47.9 | 41.1 | 60.7 | 18.3 | 23.3 | 11.0 | 12.8 | 5.0 | 62.6 | 30.6 | 0.5 |
| p value | | | $\chi^2=7.578$;<br>$p=0.0059$ | $\chi^2=0.855$;<br>$p=0.3551$ | $\chi^2=19.100$;<br>$p<0.0001$ | $\chi^2=6.828$;<br>$p=0.0090$ | $\chi^2=0.003$;<br>$p=0.9536$ | $\chi^2=11.331$;<br>$p=0.0008$ | $\chi^2=5.099$;<br>$p=0.0239$ | $\chi^2=11.843$;<br>$p=0.0006$ | $\chi^2=0.655$;<br>$p=0.4184$ | $\chi^2=0.458$;<br>$p=0.4986$ | $\chi^2=1.784$;<br>$p=0.1816$ | $\chi^2=1.340$;<br>$p=0.2471$ | $\chi^2=0.827$;<br>$p=0.3632$ |
| Place of<br>residence | City | N | 67 | 67 | 74 | 49 | 77 | 38 | 36 | 8 | 15 | 4 | 65 | 38 | 0 |
|  |  | % | 54.9 | 54.9 | 60.7 | 40.2 | 63.1 | 31.1 | 29.5 | 6.6 | 12.3 | 3.3 | 53.3 | 31.1 | 0.0 |
|  | Village | N | 205 | 139 | 195 | 112 | 203 | 79 | 94 | 23 | 38 | 16 | 208 | 115 | 4 |
|  |  | % | 60.7 | 41.1 | 57.7 | 33.1 | 60.1 | 23.4 | 27.8 | 6.8 | 11.2 | 4.7 | 61.5 | 34.0 | 1.2 |
| p value | | | $\chi^2=1.219$;<br>$p=0.2695$ | $\chi^2=6.898$;<br>$p=0.0086$ | $\chi^2=0.324$;<br>$p=0.5691$ | $\chi^2=1.946$;<br>$p=0.1630$ | $\chi^2=0.351$;<br>$p=0.5533$ | $\chi^2=2.857$;<br>$p=0.0910$ | $\chi^2=0.127$;<br>$p=0.7211$ | $\chi^2=0.009$;<br>$p=0.9256$ | $\chi^2=0.097$;<br>$p=0.7550$ | $\chi^2=0.456$;<br>$p=0.4993$ | $\chi^2=2.535$;<br>$p=0.1114$ | $\chi^2=0.334$;<br>$p=0.5633$ | $\chi^2=1.456$;<br>$p=0.2275$ |

#### Ways of using a smartphone

Young people most often used the smartphone to send messages (N = 354; 77.0%), receive and make calls (N = 339; 73.7%), and for social media (N = 322; 70, 0%). 55.0% of students (N = 253) listened to music, 49.8% (N = 229) sent and received text messages, and 46.1% of participants (N = 212) used a calculator, watch or alarm clock. 41.1% of young people (N = 189) used the smartphone for taking photos and video recordings. Adolescents used the smartphone less often to browse websites (N = 128; 27.8%), use the calendar, planner (N = 72; 15.7%), play games (N = 60; 13.0 %) or receive e-mails (N = 44; 9.6%). Very rarely, the device was used for navigation, maps (N = 21; 4.6%), reading e-books (N = 14; 3.0%) or for other purposes (N = 4; 0.9%).

Table 4 shows that girls took significantly more photos and video recordings (47.6% vs. 34.9%; *χ^2^* = 7.613; *p* = 0.0058) and used a calculator, watch and alarm clock significantly more often (51.6% vs. 40.9%; *χ^2^* = 5.301; *p* = 0.0213) than boys. Boys in turn were significantly more likely than girls to use their smartphones for playing games (18.3% vs. 7.6%; *χ^2^* = 11.695; *p* = 0.0006). Respondents aged 18-20 used a calendar more often than others (22.1%; *χ^2^* = 6.277; *p* = 0.0433). At the same time, the same age group used a smartphone less often than others to browse websites (19.8%; *χ^2^* = 10.956; *p* = 0.0042) or listen to music (41.2%; *χ^2^* = 14.104; *p* = 0.0009). It was noticed that the youngest group of respondents used messengers the least often among all respondents (69.8%; *χ^2^* = 6.657; *p* = 0.0358). The analysis showed that students of technical schools paid attention to the use of calendars (21.0% vs, 10.8%; *χ^2^* = 9.071; *p* = 0.0026), and navigation and maps (6.8% vs. 2.5%; *χ^2^* = 5.005; *p* = 0.0253) more often than high school students. High school students in turn paid more attention to using social media (75.1% vs. 64.4%; *χ^2^* = 6.279; *p* = 0.0122), taking photos and videos (12.3% vs. 7.1%; *χ^2^* = 5.168; *p* = 0.0230), and listening to music (59.3% vs. 50.2%; *χ^2^* = 3.846; *p* = 0.0499). Analyses also showed that the possibility of using social media via smartphones was more popular among rural residents (73.1%; *χ^2^* = 5.746; *p* = 0.0165).

**Table 4.** Ways of using the smartphone and sociodemographic variables.

|  |  |  | receiving and making calls | sending and receiving text messages | using a calendar | using a calculator, watch, alarm | using messengers | social media | taking photos, videos | receiving e-mail | web browsing | reading e-books | listening to music | navigation, maps | games | other |
| --- | --- | --- | --- | --- | --- | --- | --- | --- | --- | --- | --- | --- | --- | --- | --- | --- |
| Gender | Female | N | 163 | 122 | 39 | 116 | 173 | 159 | 107 | 17 | 59 | 8 | 122 | 6 | 17 | 1 |
|  |  | % | 72.4 | 54,2 | 17.3 | 51.6 | 76.9 | 70.7 | 47.6 | 7.6 | 26.2 | 3.6 | 54.2 | 2.7 | 7.6 | 0.4 |
|  | Male | N | 176 | 107 | 33 | 96 | 181 | 163 | 82 | 27 | 69 | 6 | 131 | 15 | 43 | 2 |
|  |  | % | 74.9 | 45,5 | 14.0 | 40.9 | 77.0 | 69.4 | 34.9 | 11.5 | 29.4 | 2.6 | 55.7 | 6.4 | 18.3 | 0.9 |
| p value | | | $\chi^2=0.356$;<br>$p=0.5509$ | $\chi^2=3.472$;<br>$p=0.0624$ | $\chi^2=0.943$;<br>$p=0.3315$ | $\chi^2=5.301$;<br>$p=0.0213$ | $\chi^2=0.001$;<br>$p=0.9731$ | $\chi^2=0.093$;<br>$p=0.7601$ | $\chi^2=7.613$;<br>$p=0.0058$ | $\chi^2=2.056$;<br>$p=0.1516$ | $\chi^2=0.564$;<br>$p=0.4526$ | $\chi^2=0.391$;<br>$p=0.5316$ | $\chi^2=0.108$;<br>$p=0.7428$ | $\chi^2=3.644$;<br>$p=0.0563$ | $\chi^2=11.695$;<br>$p=0.0006$ | $\chi^2=0.000$;<br>$p=1.0000$ |
| Age | 16 yrs | N | 95 | 62 | 19 | 57 | 90 | 85 | 52 | 13 | 49 | 6 | 77 | 2 | 17 | 2 |
|  |  | % | 73.6 | 48.1 | 14.7 | 44.2 | 69.8 | 65.9 | 40.3 | 10.1 | 38.0 | 4.7 | 59.7 | 1.6 | 13.2 | 1.6 |
|  | 17 yrs | N | 150 | 99 | 24 | 90 | 164 | 148 | 76 | 14 | 53 | 4 | 122 | 11 | 27 | 0 |
|  |  | % | 75.0 | 49.5 | 12.0 | 45.0 | 82.0 | 74.0 | 38.0 | 7.0 | 26.5 | 2.0 | 61.0 | 5.5 | 13.5 | 0.0 |
|  | 18-20 yrs | N | 94 | 68 | 29 | 65 | 100 | 89 | 61 | 17 | 26 | 4 | 54 | 8 | 16 | 1 |
|  |  | % | 71.8 | 51.9 | 22.1 | 49.6 | 76.3 | 67.9 | 46.6 | 13.0 | 19.8 | 3.1 | 41.2 | 6.1 | 12.2 | 0.8 |
| p value | | | $\chi^2=0.430$;<br>$p=0.8065$ | $\chi^2=0.396$;<br>$p=0.8204$ | $\chi^2=6.277$;<br>$p=0.0433$ | $\chi^2=0.940$;<br>$p=0.6249$ | $\chi^2=6.657$;<br>$p=0.0358$ | $\chi^2=2.826$;<br>$p=0.2434$ | $\chi^2=2.444$;<br>$p=0.2947$ | $\chi^2=3.323$;<br>$p=0.1898$ | $\chi^2=10.956$;<br>$p=0.0042$ | $\chi^2=1.868$;<br>$p=0.3930$ | $\chi^2=14.104$;<br>$p=0.0009$ | $\chi^2=3.807$;<br>$p=0.1490$ | $\chi^2=0.118$;<br>$p=0.9425$ | $\chi^2=2.944$;<br>$p=0.2294$ |
| School | High school | N | 178 | 114 | 26 | 110 | 190 | 181 | 111 | 27 | 74 | 6 | 143 | 6 | 27 | 3 |
|  |  | % | 73.9 | 47.3 | 10.8 | 45.6 | 78.8 | 75.1 | 46.1 | 12.3 | 30.7 | 2.5 | 59.3 | 2.5 | 11.2 | 1.2 |
|  | Tech school | N | 161 | 115 | 46 | 102 | 164 | 141 | 78 | 17 | 54 | 8 | 110 | 15 | 33 | 0 |
|  |  | % | 73.5 | 52.5 | 21.0 | 46.6 | 74.9 | 64.4 | 35.6 | 7.1 | 24.7 | 3.7 | 50.2 | 6.8 | 15.1 | 0.0 |
| p value | | | $\chi^2=0.007$;<br>$p=0.9335$ | $\chi^2=1.245$;<br>$p=0.2645$ | $\chi^2=9.071$;<br>$p=0.0026$ | $\chi^2=0.04$;<br>$p=0.8412$ | $\chi^2=1.011$;<br>$p=0.3147$ | $\chi^2=6.279$;<br>$p=0.0122$ | $\chi^2=5.168$;<br>$p=0.023$ | $\chi^2=3.691$;<br>$p=0.0547$ | $\chi^2=2.09$;<br>$p=0.1483$ | $\chi^2=0.526$;<br>$p=0.4682$ | $\chi^2=3.846$;<br>$p=0.0499$ | $\chi^2=5.005$;<br>$p=0.0253$ | $\chi^2=1.511$;<br>$p=0.2189$ | $\chi^2=2.744$;<br>$p=0.0976$ |
| Place of residence | City | N | 85 | 60 | 20 | 56 | 87 | 75 | 50 | 15 | 36 | 4 | 68 | 8 | 15 | 2 |
|  |  | % | 69,7 | 49.2 | 16.4 | 45.9 | 71.3 | 61.5 | 41.0 | 12.3 | 29.5 | 3.3 | 55.7 | 6.6 | 12.3 | 1.6 |
|  | Village | N | 254 | 169 | 52 | 156 | 267 | 247 | 139 | 29 | 92 | 10 | 185 | 13 | 45 | 1 |
|  |  | % | 75,1 | 50.0 | 15.4 | 46.2 | 79.0 | 73.1 | 41.1 | 8.6 | 27.2 | 3.0 | 54.7 | 3.8 | 13.3 | 0.3 |
| p value | | | $\chi^2=1,387$;<br>$p=0,239$ | $\chi^2=0.024$;<br>$p=0.8767$ | $\chi^2=0.069$;<br>$p=0.7926$ | $\chi^2=0.002$;<br>$p=0.9618$ | $\chi^2=2.984$;<br>$p=0.0841$ | $\chi^2=5.746$;<br>$p=0.0165$ | $\chi^2=0.001$;<br>$p=0.9784$ | $\chi^2=1.43$;<br>$p=0.2317$ | $\chi^2=0.234$;<br>$p=0.6286$ | $\chi^2=0.031$;<br>$p=0.86$ | $\chi^2=0.037$;<br>$p=0.8485$ | $\chi^2=1.512$;<br>$p=0.2188$ | $\chi^2=0.082$;<br>$p=0.7746$ | $\chi^2=2.497$;<br>$p=0.114$ |

#### Model of smartphone user

Logistic regression model using forward stepwise method (likelihood ratio) was used to characterise a typical smartphone user in this study. It was shown that smartphone users were more than 5 times more likely to be girls (OR = 5.80; *p* < 0.0001), sixteen-year-olds (OR = 0,41; *p* = 0.0456), and almost 3 times more likely to be students of technical schools (OR = 2.66; *p* = 0.0025) – Table 5.

**Table 5.** Logistic regression model characterising a smartphone user.

|  | **B** | **SE** | **Wald** | **df** | ***p*** | **OR (95% CI)** |
| --- | --- | --- | --- | --- | --- | --- |
| **Gender (1-M; 2-F)** | -1.76 | 0.37 | 22.71 | 1 | < 0.0001 | 5.80 (2.81-11.95) |
| **Age (1-16 yrs; 2-17 yrs)** | -0.88 | 0.44 | 4.00 | 1 | 0.0456 | 0.41 (0.17-0.98) |
| **Type of school (1-High; 2-Technical)** | 0.98 | 0.32 | 9.16 | 1 | 0.0025 | 2.66 (1.41-5.02) |
| **Place of residence** | -0.12 | 0.40 | 0.09 | 1 | 0.7693 | 0.89 (0.41-1.95) |
| **Living conditions** | -0.53 | 0.51 | 1.12 | 1 | 0.2908 | 0.59 (0.22-1.58) |
| **Financial situation** | 0.56 | 0.31 | 3.32 | 1 | 0.0686 | 1.75 (0.96-3.21) |

### Smartphone addiction

#### Perceiving yourself as a smartphone addict

Table 6 shows that 9.8% of students (N = 45) definitely did not perceive themselves as addicted to smartphones, and 38.3% of respondents (N = 176) were not addicted. A group of 30.2% of surveyed youth (N = 139) could not assess whether they were addicted to using a smartphone. 16.7% of respondents (N = 77) were addicted, and 5.0% of students (N = 23) were definitely addicted to using a smartphone. Girls and boys (*p* = 0.1171) and different age groups (*p* = 0.2280) perceived themselves as addicted to a smartphone equally often.

**Table 6.** Perceiving yourself as a person addicted to a smartphone and gender and age.

|  |  |  | **Gender** |  | **Age** |  |  | **Total** |
| --- | --- | --- | --- | --- | --- | --- | --- | --- |
|  |  |  | **Female** | **Male** | **16 yrs** | **17 yrs** | **18-20 yrs** |  |
| **Perceiving yourself as a person addicted to a smartphone** | **definitely no** | N | 15 | 30 | 10 | 20 | 15 | 45 |
|  |  | % | 6.7 | 12.8 | 7.8 | 10.0 | 11.5 | 9.8 |
|  | **no** | N | 83 | 93 | 51 | 82 | 43 | 176 |
|  |  | % | 36.9 | 39.6 | 39.5 | 41.0 | 32.8 | 38.3 |
|  | **no opinion** | N | 74 | 65 | 46 | 59 | 34 | 139 |
|  |  | % | 32.9 | 27.7 | 35.7 | 29.5 | 26.0 | 30.2 |
|  | yes | N | 43 | 34 | 18 | 30 | 29 | 77 |
|  |  | % | 19.1 | 14.5 | 14.0 | 15.0 | 22.1 | 16.7 |
|  | definitely<br>yes | N | 10 | 13 | 4 | 9 | 10 | 23 |
|  |  | % | 4.4 | 5.5 | 3.1 | 4.5 | 7.6 | 5.0 |
| Total |  | N | 225 | 235 | 129 | 200 | 131 | 460 |
|  |  | % | 100.0 | 100.0 | 100.0 | 100.0 | 100.0 | 100.0 |
| Test of independence | | | $\chi^2 = 7.380; p = 0.1171$ | | | $\chi^2 = 10.559; p = 0.2280$ | | |

#### Problems with establishing face-to-face contacts

22.2% of young people (N = 102) had problems with face-to-face relationships. 77.8% of respondents (N = 358) did not think they had this type of problem. Table 7 shows that problems with establishing social relationships more often affected students of technical schools than students of high schools (27.4% vs. 17.4%; *χ^2^* = 6.609; *p* = 0.0101). Place of residence did not significantly affect this issue.

**Table 7.** Problems with establishing face-to-face social relationships and type of school and place of residence.

|  |  |  | Type of school |  | Place of residence |  | Total |
| --- | --- | --- | --- | --- | --- | --- | --- |
|  |  |  | High school | Tech school | City | Village |  |
| Problems with establishing face-to-face social relationships | yes | N | 42 | 60 | 29 | 73 | 102 |
|  |  | % | 17.4 | 27.4 | 23.8 | 21.6 | 22.2 |
|  | no | N | 199 | 159 | 93 | 265 | 358 |
|  |  | % | 82.6 | 72.6 | 76.2 | 78.4 | 77.8 |
| Total |  | N | 241 | 219 | 122 | 338 | 460 |
|  |  | % | 100.0 | 100.0 | 100.0 | 100.0 | 100.0 |
| Test of independence | | | $\chi^2 = 6.609; p = 0.0101$ | | $\chi^2 = 0.245; p = 0.6204$ | | |

#### Neglecting home/school duties

Table 8 shows that girls significantly more often than boys admitted to neglecting household or school duties as a result of using a smartphone (61.8% vs. 51.5%; *χ^2^* = 4.951; *p* = 0.0261). More frequent cases of neglect of home/school duties also concerned respondents aged 18-20 (62.6%; *χ^2^* = 4.748; *p* = 0.0931) compared to 16-year-olds or 17-year-olds however, this relationship was not statistically significant

**Table 8.** Neglecting home/school duties as a result of smartphone use and gender and age.

|  |  |  | Gender |  | Age |  |  | Total |
| --- | --- | --- | --- | --- | --- | --- | --- | --- |
|  |  |  | Female | Male | 16 yrs | 17 yrs | 18-20 yrs |  |
| Neglecting home/school duties as a result of smartphone use | yes | N | 139 | 121 | 76 | 102 | 82 | 260 |
|  |  | % | 61.8 | 51.5 | 58.9 | 51.0 | 62.6 | 56.5 |
|  | no | N | 86 | 114 | 53 | 98 | 49 | 200 |
|  |  | % | 38.2 | 48.5 | 41.1 | 49.0 | 37.4 | 43.5 |
| Total |  | N | 225 | 235 | 129 | 200 | 131 | 460 |
|  |  | % | 100.0 | 100.0 | 100.0 | 100.0 | 100.0 | 100.0 |
| Test of independence | | | $\chi^2 = 4.951; p = 0.0261$ | | $\chi^2 = 4.748; p = 0.0931$ | | | |

#### Inability to spend time without a smartphone

Table 9 shows that in the opinion of 67.8% of respondents (N = 312) using a smartphone increases the standard of living. 32.2% of youth (N = 148) did not share this opinion. Most often, young people claimed that they could not spend a month without a smartphone because it is necessary to communicate with others (N = 245; 53.3%). The device was necessary to use the Internet for 11.5% of respondents (N = 53), and 7.6% of respondents (N = 35) believed that it made them more mobile. 6.7% of students (N = 31) could not spend a month without a smartphone due to lack of access to entertainment. Rarely did young people say that they would not spend a month without a smartphone because it helps them with their homework (N = 17; 3.7%), provides them with rest (N = 15; 3.3%), or for some other reason (N = 3; 0.7%). 13.3% of students (N = 61) would be able to spend a month without using a smartphone. Girls were more likely than boys (58.2% vs. 48.5%; *p* = 0.0028%) to be unable to spend a month without a smartphone because they considered it necessary in communicating with others. Age did not play a significant role in this aspect. High school students more often (59.3%; *p* = 0.0017) believed that they could not spend a month without a smartphone, because they needed it to communicate with others. Place of residence did not significantly differentiate this aspect.

**Table 9.**
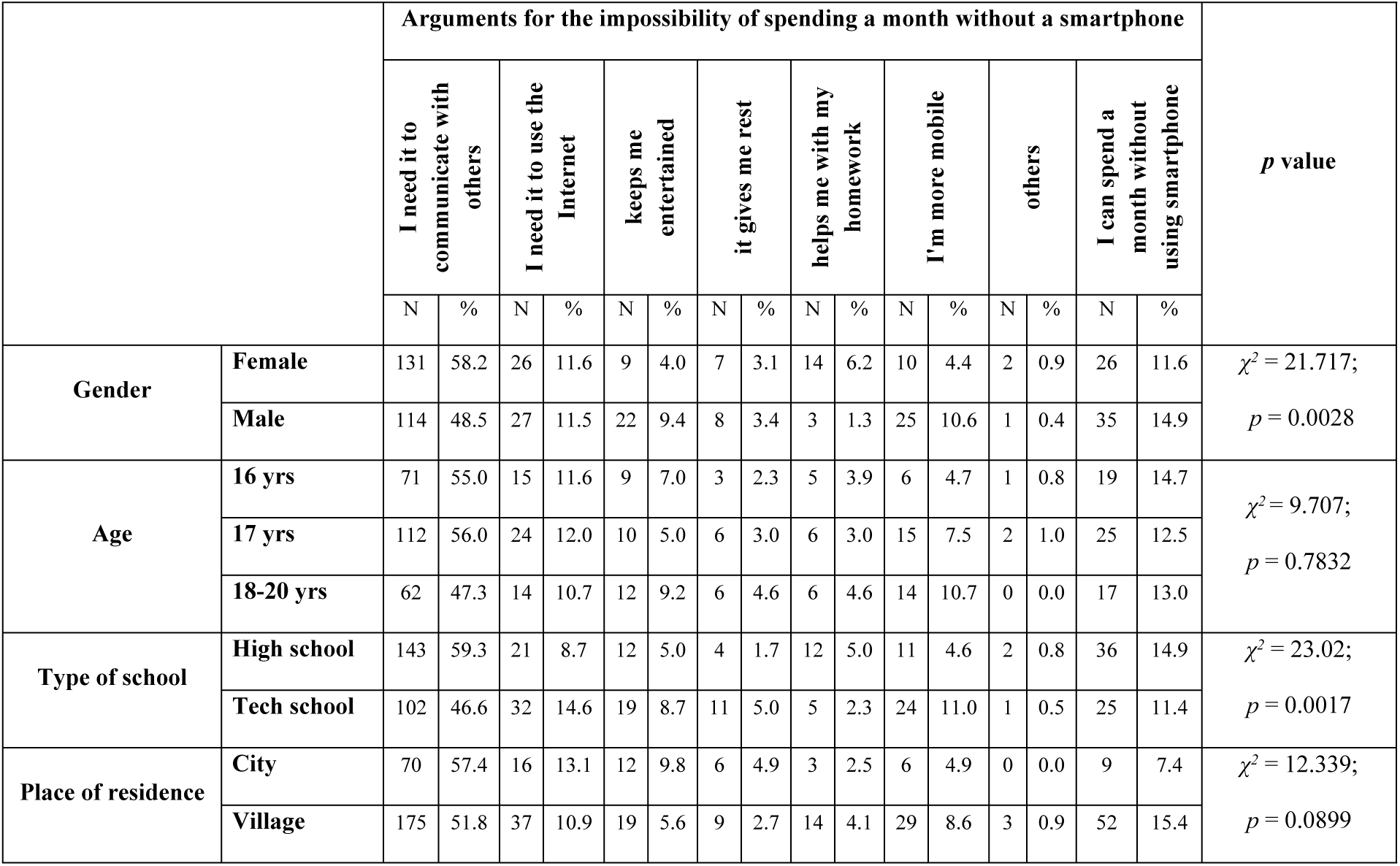
Associations between arguments for the impossibility of spending a month without a smartphone and sociodemographic variables.

#### Returning home in case forgetting the smartphone

Table 10 shows that 37.6% of youth (N = 173) would definitely come back home if they forgot to take their smartphone. Almost the same number of respondents (N = 159; 34.6%) would not return. 27.8% of respondents (N = 128) never forgot their smartphone and always had it with them. It was noticed that as the age of the respondents increased, the percentage of those who would definitely return home for their smartphone increased. However, the differences were not significant (*p* = 0.0694). There was no relationship between the discussed situation and the gender, type of school or place of residence of the respondents.

**Table 10.**
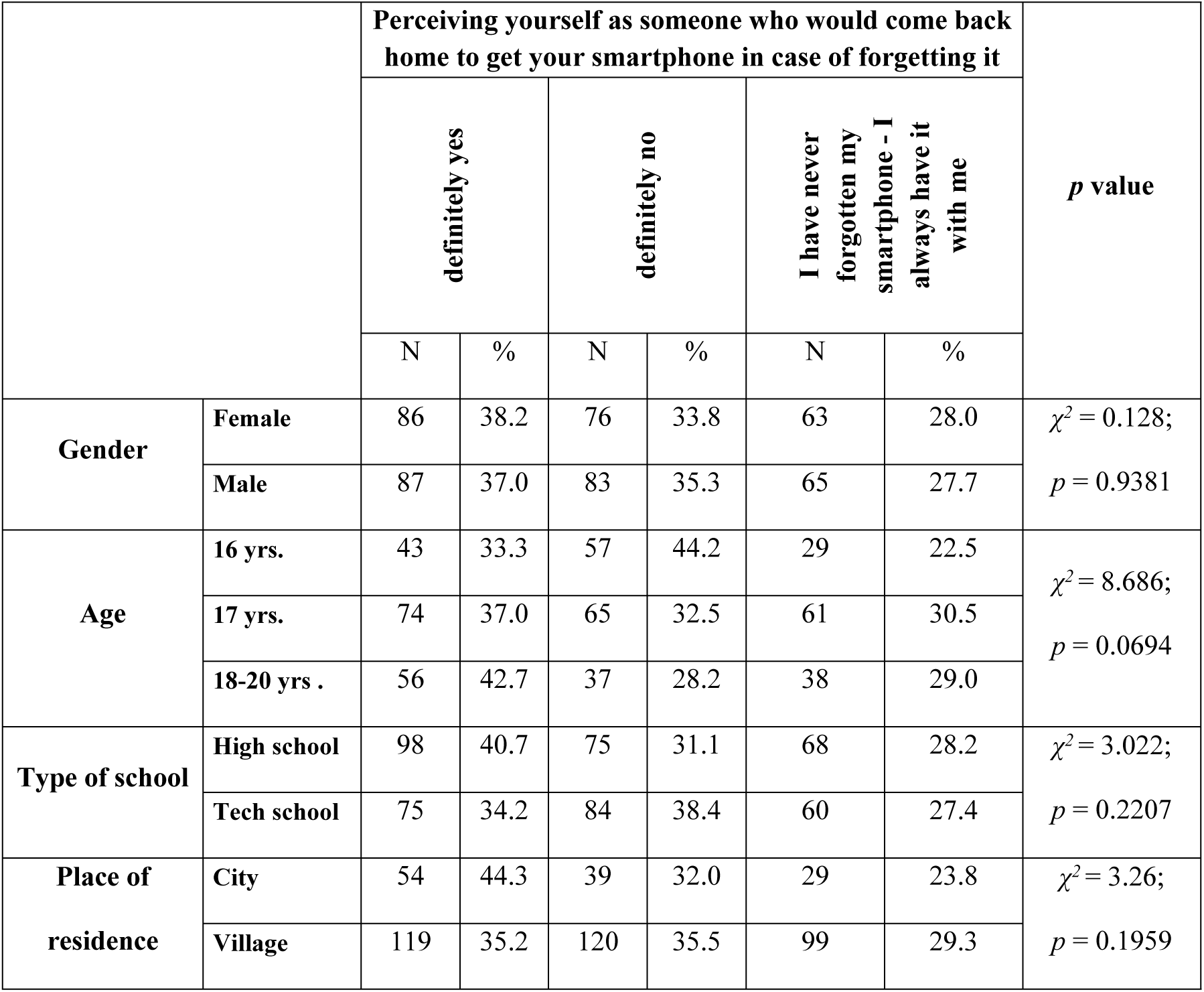
Associations between perceiving yourself as a person who would come back home to get a smartphone in case of forgetting it and sociodemographic variables.

#### Situations of not using the smartphone

Table 11 illustrates that respondents most often did not use a smartphone during meals with their families at home (N = 277; 60.2%). Almost half of the students (N = 221; 48.0%) did not use a smartphone while riding a motorcycle or bicycle, and 46.3% of respondents (N = 213) did not use the device in the cinema, theatre or at a concert. A group of 40.9% of young people (N = 188) admitted that they did not use a smartphone while dining in a restaurant with family or friends, while 37.0% of respondents (N = 170) did not use it while crossing the street. 23.3% of respondents (N = 107) did not use a smartphone during social events, and 20.4% of students (N = 94) did not use it during lessons or extracurricular activities. A group of 7.8% of the surveyed youth (N = 36) admitted that there are no situations in which they would not use a smartphone. The analysis of the collected research material allowed us to conclude that young people did not use a smartphone regardless of gender, but only in one situation depending on age (*p* < 0.0001). It turned out that respondents aged 16 (36.4%) did not use a smartphone more often during lessons or extracurricular activities compared to 17-year-olds (15.0%) or the age group 18-20 (13.0%). %). High school students did not use a smartphone while riding a motorcycle or cycling more often than students of a technical school (53.9% vs. 41.6%; *p* = 0.0079). It was also found that rural residents were more likely than urban residents (63.0% vs. 52.5%; *p* = 0.0411) not to use a smartphone during meals with their family at home.

**Table 11.** Associations between situations of not using a smartphone and sociodemographic variables.

|  |  |  | during meals with family at home | during meals in a restaurant with family or friends | at the cinema, theatre, concert | during social events | while riding a motorcycle, riding a bicycle | during lessons, extracurricular activities | when crossing the street | There are no situations when I wouldn't use my smartphone |
| --- | --- | --- | --- | --- | --- | --- | --- | --- | --- | --- |
| Gender | Female | N | 129 | 89 | 98 | 49 | 115 | 44 | 80 | 17 |
|  |  | % | 57.3 | 39.6 | 43.6 | 21.8 | 51.1 | 19.6 | 35.6 | 7.6 |
|  | Male | N | 148 | 99 | 115 | 58 | 106 | 50 | 90 | 19 |
|  |  | % | 63.0 | 42.1 | 48.9 | 24.7 | 45.1 | 21.3 | 38.3 | 8.1 |
| p value | | | $\chi^2 = 1.529$;<br>$p = 0.2162$ | $\chi^2 = 0.315$;<br>$p = 0.5748$ | $\chi^2 = 1.338$;<br>$p = 0.2473$ | $\chi^2 = 0.543$;<br>$p = 0.4613$ | $\chi^2 = 1.660$;<br>$p = 0.1976$ | $\chi^2 = 0.209$;<br>$p = 0.6472$ | $\chi^2 = 0.371$;<br>$p = 0.5424$ | $\chi^2 = 0.045$;<br>$p = 0.8326$ |
| Age | 16 yrs. | N | 77 | 49 | 67 | 27 | 67 | 47 | 50 | 6 |
|  |  | % | 59.7 | 38.0 | 51.9 | 20.9 | 51.9 | 36.4 | 38.8 | 4.7 |
|  | 17 yrs. | N | 122 | 84 | 89 | 48 | 91 | 30 | 73 | 16 |
|  |  | % | 61.0 | 42.0 | 44.5 | 24.0 | 45.5 | 15.0 | 36.5 | 8.0 |
|  | 18-20 yrs. | N | 78 | 55 | 57 | 32 | 63 | 17 | 47 | 14 |
|  |  | % | 59.5 | 42.0 | 43.5 | 24.4 | 48.1 | 13.0 | 35.9 | 10.7 |
| **p value** | | | $\chi^2 = 0.091$;<br>$p = 0.9555$ | $\chi^2 = 0.617$;<br>$p = 0.7344$ | $\chi^2 = 2.320$;<br>$p = 0.3136$ | $\chi^2 = 0.554$;<br>$p = 0.7582$ | $\chi^2 = 1.302$;<br>$p = 0.5214$ | $\chi^2 = 28.424$;<br>$p < 0.0001$ | $\chi^2 = 0.263$;<br>$p = 0.8766$ | $\chi^2 = 3.297$;<br>$p = 0.1923$ |
| **Type of school** | **High school** | N | 149 | 107 | 120 | 58 | 130 | 53 | 99 | 23 |
|  |  | % | 61.8 | 44.4 | 49.8 | 24.1 | 53.9 | 22.0 | 41.1 | 9.5 |
|  | **Tech school** | N | 128 | 81 | 93 | 49 | 91 | 41 | 71 | 13 |
|  |  | % | 58.4 | 37.0 | 42.5 | 22.4 | 41.6 | 18.7 | 32.4 | 5.9 |
| **p value** | | | $\chi^2 = 0.547$;<br>$p = 0.4597$ | $\chi^2 = 2.608$;<br>$p = 0.1063$ | $\chi^2 = 2.477$;<br>$p = 0.1155$ | $\chi^2 = 0.184$;<br>$p = 0.6679$ | $\chi^2 = 7.056$;<br>$p = 0.0079$ | $\chi^2 = 0.755$;<br>$p = 0.385$ | $\chi^2 = 3.692$;<br>$p = 0.0547$ | $\chi^2 = 2.07$;<br>$p = 0.1502$ |
| **Place of residence** | **City** | N | 64 | 55 | 62 | 31 | 62 | 21 | 46 | 12 |
|  |  | % | 52.5 | 45.1 | 50.8 | 25.4 | 50.8 | 17.2 | 37.7 | 9.8 |
|  | **Village** | N | 213 | 133 | 151 | 76 | 159 | 73 | 124 | 24 |
|  |  | % | 63.0 | 39.3 | 44.7 | 22.5 | 47.0 | 21.6 | 36.7 | 7.1 |
| **p value** | | | $\chi^2 = 4.172$;<br>$p = 0.0411$ | $\chi^2 = 1.219$;<br>$p = 0.2695$ | $\chi^2 = 1.362$;<br>$p = 0.2433$ | $\chi^2 = 0.43$;<br>$p = 0.5122$ | $\chi^2 = 0.513$;<br>$p = 0.474$ | $\chi^2 = 1.06$;<br>$p = 0.3032$ | $\chi^2 = 0.04$;<br>$p = 0.8416$ | $\chi^2 = 0.93$;<br>$p = 0.3349$ |

#### Smartphone use and health effects

Table 12 shows that most students were aware and shared the opinion that using a smartphone may negatively affect their health (N = 324; 70.4%). 10.4% of respondents (N = 48) did not notice any negative aspects of using a smartphone on health, and 19.1% of participants (N = 88) had no opinion on this subject. It was noticed that 16-year-olds more often (16.3%) than 17-year-olds (6.5%) believed that a smartphone could not negatively affect their health (*χ^2^* = 10.890; *p* = 0.0278). Gender did not play a significant role here (*χ^2^* = 2.879; *p* = 0.2371).

**Table 12.**
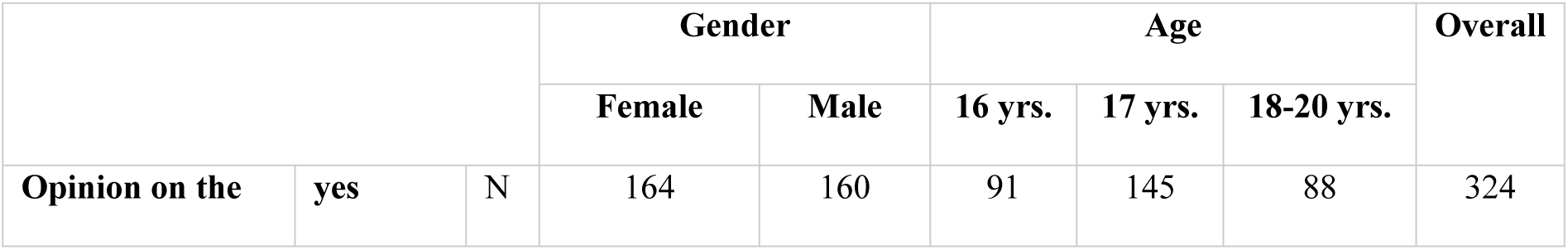
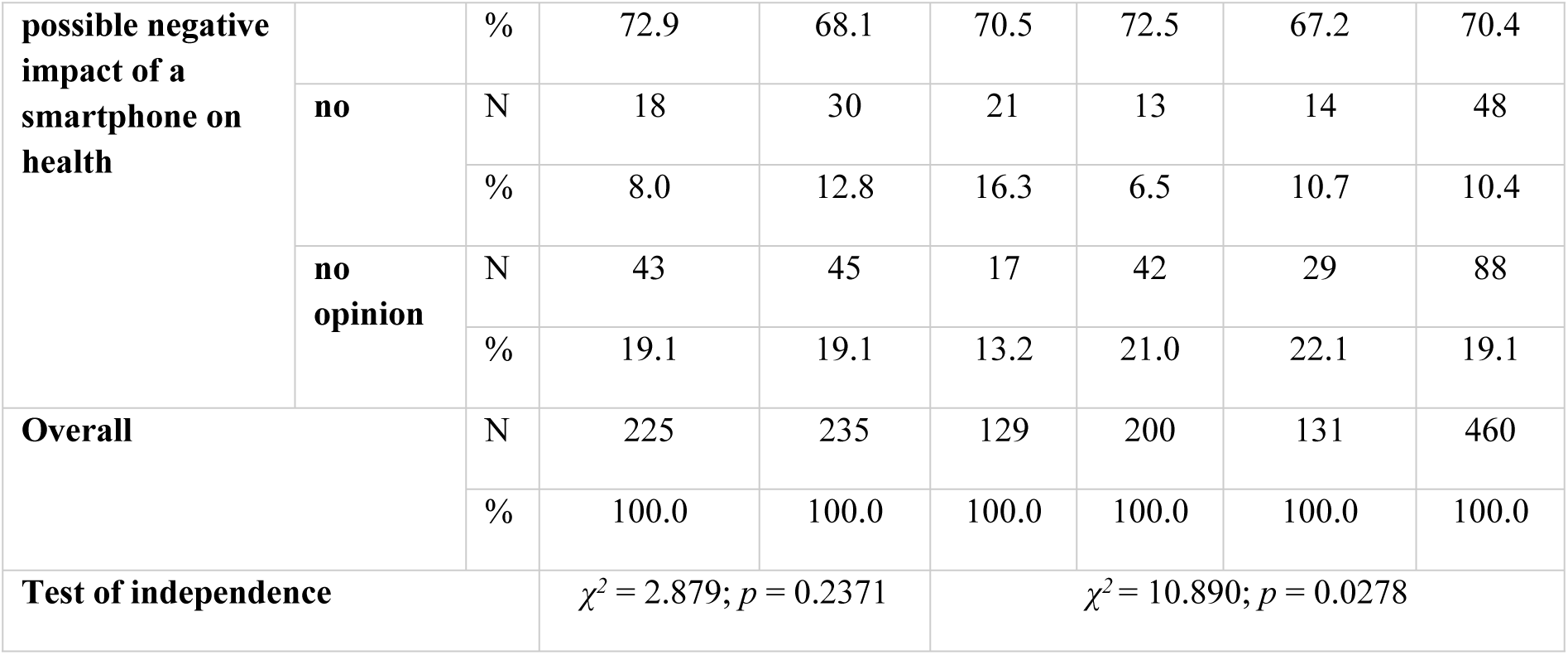
Associations between opinion on the possible negative impact of a smartphone on health and gender and age.

#### The level of smartphone addiction

The level of smartphone addiction was assessed using the KBUTK scale, in 4 dimensions and the overall score. Each dimension and the overall result included a scale of 1-5 points, with higher scores corresponding to greater addiction.

Table 13 shows that the highest level of smartphone addiction was related to “Addiction to camera functions” (3.16±0.66). The second indicator in which young people obtained the highest results was the “Need of acceptance and closeness” dimension (2.12±0.84). There was little dependence on “Indirect communication” (1.89±0.81) and “Addiction to phone calls and text messages” (1.61±0.66). The overall level of smartphone addiction was 2.17±0.58 points on a scale of 1-5 points.

**Table 13.**
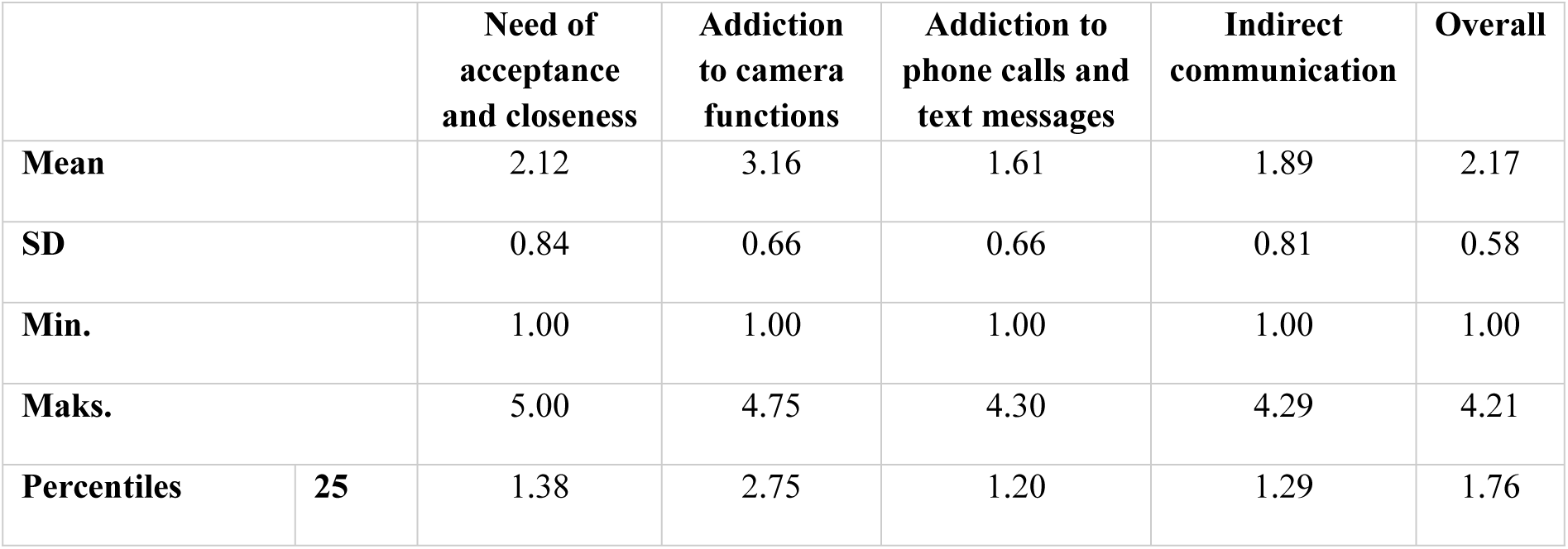
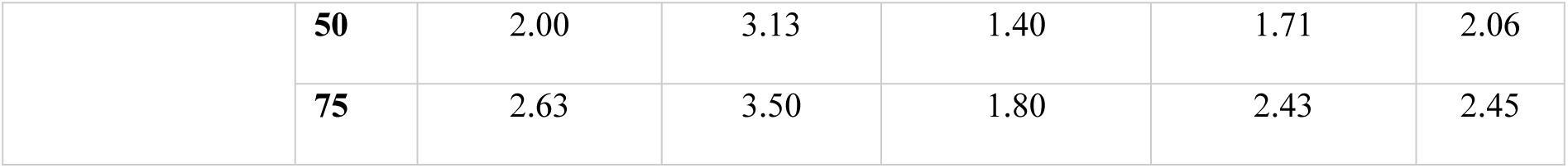
Smartphone addiction scale (KBUTK).

|  |  | **Need of acceptance and closeness** | **Addiction to camera functions** | **Addiction to phone calls and text messages** | **Indirect communication** | **Overall** |
| --- | --- | --- | --- | --- | --- | --- |
| **Mean** |  | 2.12 | 3.16 | 1.61 | 1.89 | 2.17 |
| **SD** |  | 0.84 | 0.66 | 0.66 | 0.81 | 0.58 |
| **Min.** |  | 1.00 | 1.00 | 1.00 | 1.00 | 1.00 |
| **Maks.** |  | 5.00 | 4.75 | 4.30 | 4.29 | 4.21 |
| **Percentiles** | **25** | 1.38 | 2.75 | 1.20 | 1.29 | 1.76 |
|  | **50** | 2.00 | 3.13 | 1.40 | 1.71 | 2.06 |
|  | **75** | 2.63 | 3.50 | 1.80 | 2.43 | 2.45 |

It was shown that the overall rate of smartphone addiction was significantly higher (*p* < 0.0001) among girls (2.31 pts) than boys (2.03 pts). This difference has been also manifested in four aspects of KBUTK: “Need of acceptance and closeness” (*p* < 0.0001), “Addiction to camera functions” (*p* < 0.0001), “Addiction to phone calls and text messages” (*p* = 0.0014), and “Indirect communication” (*p* < 0.0001). The level of smartphone addiction was not found to be significantly related to the age of the respondents. However, it was shown that high school students were more often addicted to the camera functions (3.26 vs. 3.04; *p* = 0.0005), and students of technical schools were more often addicted to calls and text messages (1.76 vs. 1.47; *p* = 0.0013). The place of residence of young people did not significantly affect the level of smartphone addiction, both in general and in individual aspects – Table 14.

**Table 14.**
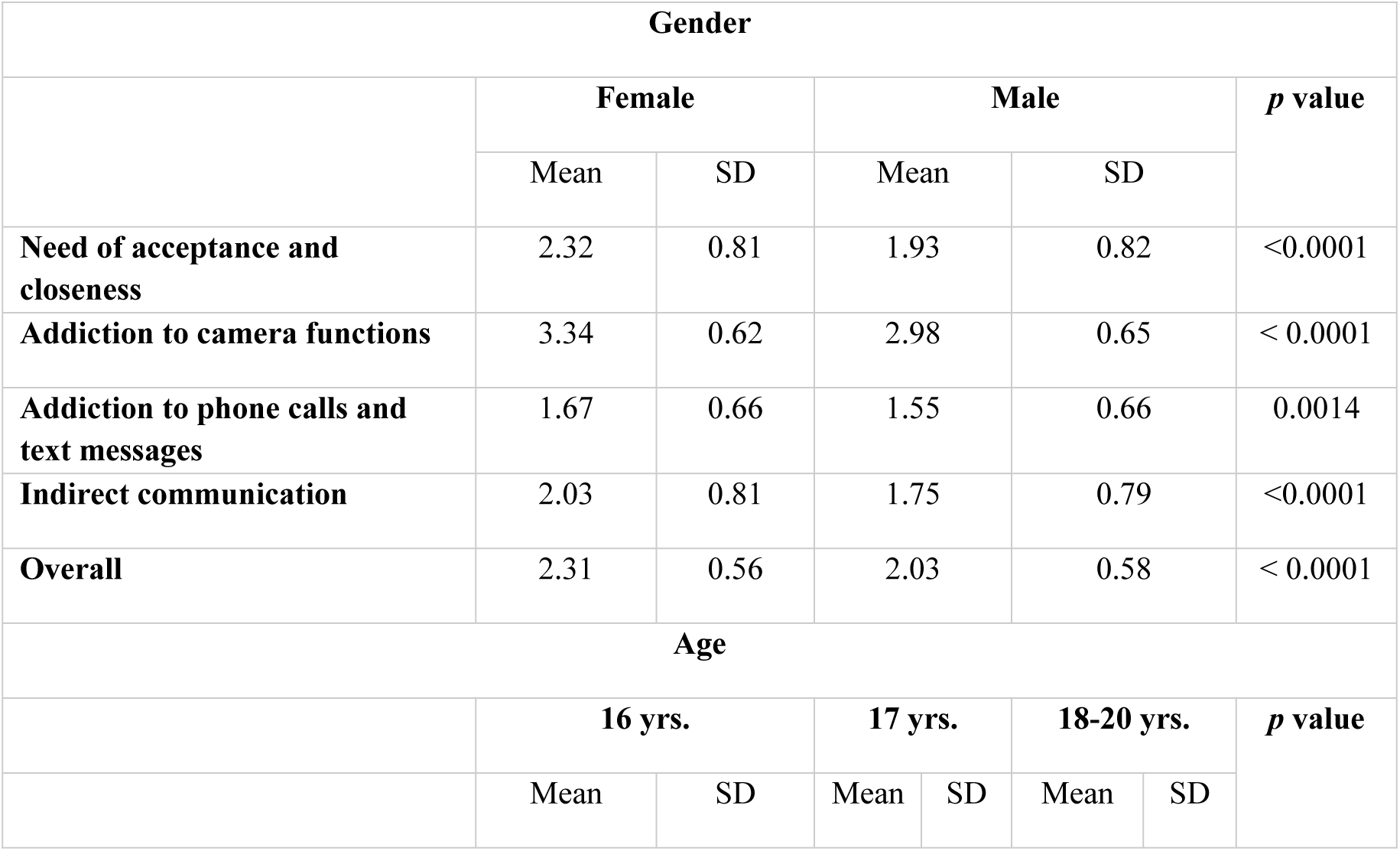
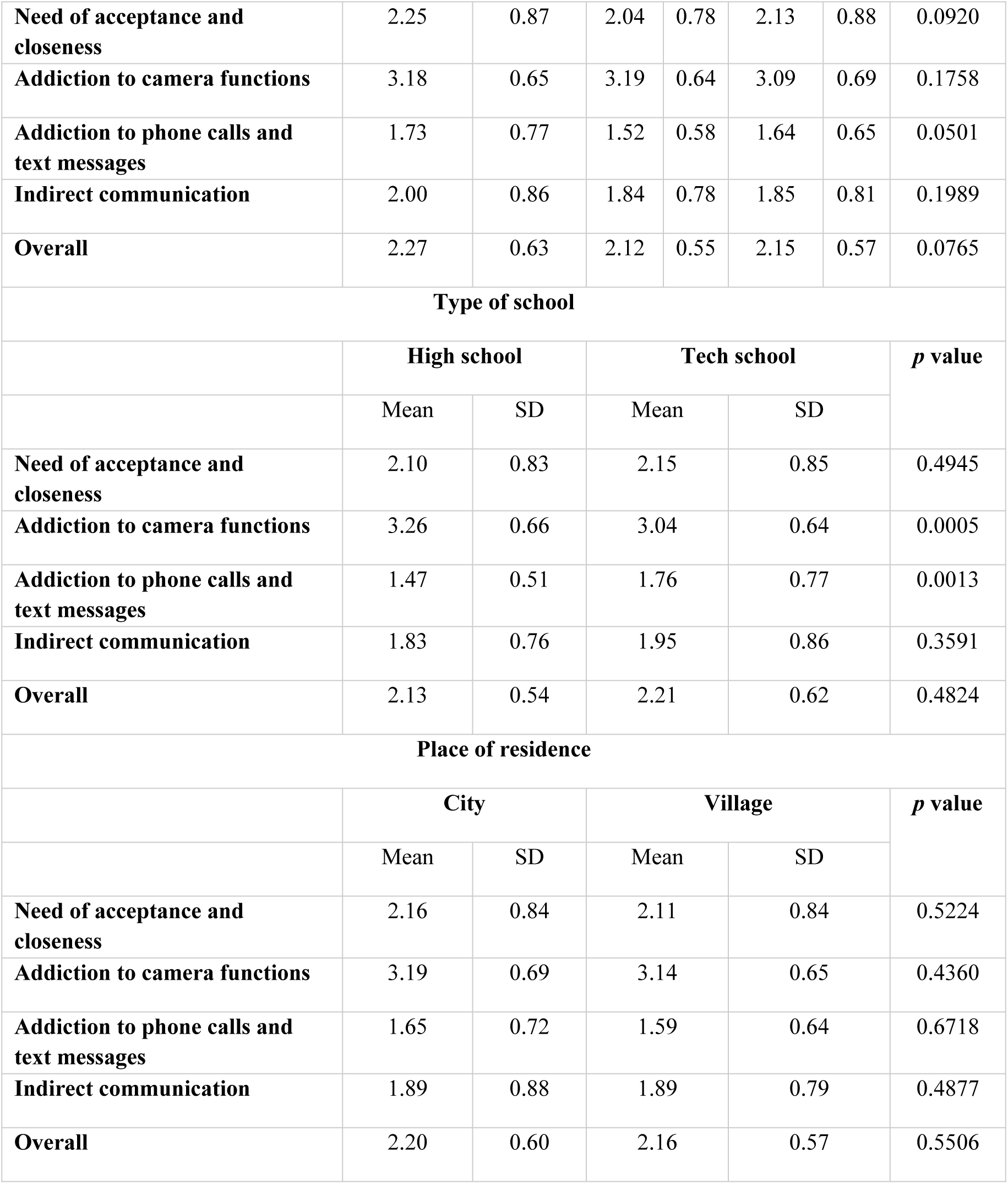
Associations between smartphone addiction scale and sociodemographic variables.

#### Perceiving oneself as a person addicted to a smartphone and KBUTK results

Table 15 shows that perceiving oneself as a smartphone addict was significantly related to the results of KBUTK scale. It was found that higher scores on “Need of acceptance and closeness” were obtained by students who perceived themselves as definitely addicted to the smartphone (2.57 pts; *p* = 0.0002). Similarly, respondents who perceive themselves as definitely addicted to a smartphone obtained significantly higher scores on the scale: “Addiction to camera functions” (3.42 pts; *p* = 0.0001) and “Addiction to phone calls and text messages” (1.82 pts; *p* < 0, 0001). Respondents who negatively assessed the possibility of smartphone addiction had a significantly lower actual level of addiction in the field of “Indirect communication” (1.75 pts; *p* = 0.0070). The general level of phone addiction correlated positively with the perception of oneself as a smartphone addict (*rho* = 0.223; *p* < 0.0001). The results indicate that smartphone addiction is objective.

**Table 15.** Perceiving yourself as a person addicted to a smartphone and actual addiction (KBUKT).

| Perceiving yourself as a person addicted to a smartphone |  | Need of acceptance and closeness | Addiction to camera functions | Addiction to phone calls and text messages | Indirect communication | Overall |
| --- | --- | --- | --- | --- | --- | --- |
| definitely no | Mean | 1.80 | 2.79 | 1.56 | 1.75 | 1.96 |
|  | SD | 0.81 | 0.66 | 0.88 | 0.85 | 0.69 |
| no | Mean | 1.98 | 3.08 | 1.51 | 1.75 | 2.05 |
|  | SD | 0.71 | 0.60 | 0.60 | 0.70 | 0.46 |
| no opinion | Mean | 2.29 | 3.28 | 1.70 | 2.09 | 2.31 |
|  | SD | 0.88 | 0.68 | 0.70 | 0.90 | 0.65 |
| yes | Mean | 2.21 | 3.26 | 1.64 | 1.90 | 2.23 |
|  | SD | 0.85 | 0.64 | 0.53 | 0.80 | 0.51 |
| definitely yes | Mean | 2.57 | 3.42 | 1.82 | 1.96 | 2.42 |
|  | SD | 1.08 | 0.69 | 0.72 | 0.82 | 0.65 |
| Overall | Mean | 2.12 | 3.16 | 1.61 | 1.89 | 2.17 |
|  | SD | 0.84 | 0.66 | 0.66 | 0.81 | 0.58 |
| $\rho$ | | 0.198 | 0.205 | 0.233 | 0.132 | 0.223 |
| $p$ value | | 0.0002 | 0.0001 | <0.0001 | 0.0070 | <0.0001 |

#### General level of smartphone addiction and sociodemographic variables

We examined how all sociodemographic variables (gender, age, type of school, place of residence, living conditions, financial situation, parents’ education) had an impact on the overall level of smartphone addiction. For this purpose, the general level of smartphone addiction was divided into two groups: no addiction (coded: 0), and risk of addiction/addiction (coded: 1). The impact of selected variables on smartphone addiction was checked using a multiple logistic regression analysis.

Logistic regression model using input method shows that smartphone addiction was significantly influenced by: gender (*p* < 0.0001), living conditions (*p* = 0.0049), and type of school (*p* = 0.0021). Smartphone addiction was found more often among respondents living in apartment blocks (OR = 2.63; 1.34-5.16), students of technical schools (OR = 2.05; 1.30-3.23), and almost 3 times less frequently (OR=0.36; 0.23-0.56) in boys than in girls. The forward selection model confirmed the significant impact of the three previously indicated variables on the occurrence of smartphone addiction: gender (OR = 0.35; 0.22-0.55), living conditions (OR = 2.08; 1.17-3.70), and school type (OR = 2.00; 1.28-3.13). Smartphone addiction was significantly more common among girls, students of technical schools, and respondents living in blocks of flats - Table 16.

**Table 16.** Association between smartphone addiction and sociodemographic variables.

| Logistic regression model |  | B | SE | *p* | OR | 95% CI (OR) |  |
| --- | --- | --- | --- | --- | --- | --- | --- |
|  |  |  |  |  |  | lower | upper |
| input | Gender | -1.03 | 0.23 | < 0.0001 | 0.36 | 0.23 | 0.56 |
|  | Age | -0.20 | 0.14 | 0.1408 | 0.82 | 0.63 | 1.07 |
|  | Place of residence | 0.26 | 0.27 | 0.3470 | 1.29 | 0.76 | 2.20 |
|  | Living conditions | 0.97 | 0.34 | 0.0049 | 2.63 | 1.34 | 5.16 |
|  | **Type of school** | 0.72 | 0.23 | 0.0021 | 2.05 | 1.30 | 3.23 |
| **forward selection** | **Gender** | -1.05 | 0.23 | 0.0000 | 0.35 | 0.22 | 0.55 |
|  | **Living conditions** | 0.73 | 0.29 | 0.0130 | 2.08 | 1.17 | 3.70 |
|  | **Type of school** | 0.69 | 0.23 | 0.0022 | 2.00 | 1.28 | 3.13 |

## Discussion

The aim of this study was to examined the prevalence of smartphone addiction among Polish secondary school students, and identified factors related to such phenomenon, compared to the respondents’ subjective opinions about the usage, and possible addiction with an objective assessment of the degree of smartphone addiction. Therefore, we have carried out demographic characteristics of smartphones users, as well as the main features that determine the purchase of device, and ways to use it by adolescents.

Our study revealed that 100% of respondents use a smartphone, and over 96% of them own such a device. It was also shown that as many as 94.8% of the surveyed youth connect to the Internet via a smartphone. Our findings are consistent with previous research that has shown high levels of smartphone ownership among high school students from wide geographical regions [11, 26–29]. In this study, female students were more than 5 times more likely to be smartphone users which is consistent with previous results [30, 31]. However, the more detailed results of these studies are interesting. We have found that boys and girls use phones for different reasons: girls spend more time on social media or texting, while boys are more interested in video games, media sharing, and Internet searches [32].

We have asked participants if they perceived themselves as an smartphone addict. Students were also asked to describe how mobile device affects their everyday routine, including: problems with establishing face-to-face contacts, neglecting home/school duties, or inability to spend time without a smartphone. Our research has shown that 5% of adolescents considered themselves addicted to smartphones, while less than 17% of respondents found themselves at risk of addiction. More than 30% of respondents had no opinion on this subject. Our results also allowed us to determine that over 22% of the surveyed youth noticed problems with face-to-face relationships. Moreover, respondents fell asleep with phone and used it at night. More than half of participants addicted to and at risk of addiction neglected their home or school duties. According to nationwide report conducted on 22,086 students aged 12-18, almost half (49%) of teenagers try to always be “on call” [33]. Slightly fewer (45.8%) teenagers try to have a phone with them at all times [33]. What may be disturbing is the fact that a high percentage (76.5%) of participant in our study are teenagers carrying a smartphone with them all the time. Moreover, research highlights disturbing phenomenon that may indicate smartphone abuse or lead to addiction. It should be noted that having a phone close gives a sense of security to over 47% of respondents [33]. Less than 46% of the surveyed youth ensures that they always have their device with them, even while sleeping [33]. In our study over 59% of respondents admitted that they use the phone at night, and 72% of them put the phone next to the bed at night which is consistent with the discussed report. Moreover, Dębski’s research shows that over 37% of students cannot imagine their daily life without a mobile phone, and almost 27% of respondents would return for it if they forgot to take their mobile phone with them [33]. Less than 14% of students in our research declare that they would be able to spend a month without a smartphone. Among the arguments for such an impossibility, respondents claimed that they needed a phone to communicate with others (53.3%), and to connect to the Internet (11.5%). As many as 27.8% of respondents in our study have never forgotten to take their smartphone with them, and 37.6% of students would return for it if it happened. Research conducted in Spain indicates that 20% of young people aged 13-20 use their mobile devices incorrectly [34]. Similarly, research conducted in Great Britain has shown that 10% of young people aged 11-18 use their phones in an improperly way [35]. A survey conducted in Italy among youth aged 11 to 18 shows that teenagers unlock their phones 75 to 120 times a day. Moreover, as many as 83% of Italian youth spend 4 hours a day on social media, which is two months of uninterrupted use per year [36]. The results of our own research showed that over 44% of respondents were unable to specify how many times a day they use their phone because they claimed that they use their smartphone all the time.

The self-assessment of addiction corresponds with the respondents’ actual smartphone addiction in our study, because the analysis of the KBUTK test confirmed such addiction in 6.1% of respondents. A large group of respondents (32.2%) were at risk of addiction. 61.7% of respondents showed no addiction to smartphones. Polish reports showed that less than 3% of the surveyed young people show symptoms of addictive use of smartphones [33]. Similarly, research conducted among 470 secondary school students in western Poland showed that less than 4% of young people are addicted to mobile phones. Unfortunately, nearly 35% of respondents were at risk of addiction [37]. Our research confirmed that addiction or the risk of addiction to smartphones is 3 times more common in females than in males, which is convergent with the results of a systematic review amongst children and young people [6]. Moreover, such addiction was correlated with communication and networking applications similarly to our analyses. The above study also showed that the risk of addiction increases significantly with the age of the respondents, but in our research we did not found such a relationship [6]. Meanwhile, similarly to Warzecha and Pawlak, living conditions and school type were found to be significant predictors of mobile device addiction [37]. In turn, out of 248 junior high school students, 2% were addicted to smartphones. It should be noted here that these addicted respondents were girls [38]. These studies also showed that almost 30% of respondents were at risk of addiction. Females were again a larger group at risk of developing addiction than males. Less than 70% of respondents were not at risk of developing mobile phone addiction [38]. Another study using KBUTK conducted among Polish adolescents aged 13 to 19 again confirmed that there are more girls than boys in the group of respondents addicted to smartphones [39]. Less than 4% of girls and 0.3% of boys met the criteria for smartphone addiction. The group at risk of developing addiction included 23.4% of girls and 12% of boys [39].

## Conclusion

This study concluded that a typical Polish smartphone user during adolescence was more than 5 times more likely to be a girl and almost 3 times more likely to be a student of technical secondary school. Females were also more likely than males to have a smartphone addiction. What is symptomatic, the general level of smartphone addiction correlated positively with the perception of being a smartphone addict. It was also found that the overuse of smartphones leads to problematic behaviors resulting in: problems with face-to-face contacts, avoiding school or home duties, or carrying the device all the time even at night. This study demonstrated that smartphone usage contains potential risk in a group of adolescents and its negative effects should attract the attention of parents, teachers and students alike.

## Data Availability

The data underlying the results presented in the study are available from Communities in Repository UR https://repozytorium.ur.edu.pl/communities/e0ac40a9-69ba-4866-8420-ed2464213d1b

## Acknowledgments

Authors would like to thank the school authorities and the anonymous participants of this study.

## Notes

### Competing Interest Statement

The authors have declared no competing interest.

### Funding Statement

The author(s) received no specific funding for this work.

### Author Declarations

The Bioethics Committee of the Rzeszow University – Resolution No. 28/02/2019

